# Apathy, intentions, explicit attitudes, and approach-avoidance tendencies in physical activity behavior

**DOI:** 10.1101/2024.07.16.24310493

**Authors:** Ata Farajzadeh, François Jabouille, Nickolas Benoit, Olivier Bezeau, Tristan Bourgie, Benjamin Gerro, Jacob Ouimet, Matthieu P. Boisgontier

## Abstract

**Background:** Greater behavioral apathy has been shown to be associated with lower engagement in physical activity. However, the mechanisms underlying this association remain overlooked and poorly understood. Intentions, explicit attitudes, and approach-avoidance tendencies toward physical activity may play a central role in the relationship, given their strong links to motivation and physical activity.

**Methods:** An online study was conducted in 365 participants aged 54±18 years. All measures were assessed using questionnaires, except approach-avoidance tendencies, which were derived from reaction times in an approach-avoidance task. Component mediation analyses based on multiple linear regressions were conducted to examine whether the intention to be physically active mediated the relationship between behavioral apathy levels and usual physical activity levels, and whether explicit attitudes and approach-avoidance tendencies mediated the relationship between behavioral apathy and the intention.

**Results:** Results showed that weaker intentions to be physically active mediated the association between higher behavioral apathy and lower usual weekly levels of moderate-to-vigorous physical activity. In addition, explicit attitudes mediated the effect of behavioral apathy on intentions to be physically active. Results on approach-avoidance tendencies showed a significant three-way interaction between apathy, stimulus (physical activity vs. sedentary behavior), and action direction (approach vs. avoidance) on corrected reaction time (b = 19.6; 95CI = 2.0 to 37.3; p = .029), with higher apathy being associated with a greater tendency to avoid physical activity stimuli and to approach sedentary stimuli. However, we found no evidence suggesting that these tendencies mediated the effect of apathy on intentions or habitual physical activity. Based on our data, a mean item score greater than 34.5% of the scale range (e.g., >3.07 on a 1–7 scale) is indicative of behavioral apathy.

**Conclusion:** This study provides new insights into the role of intentions, explicit attitudes, and approach-avoidance tendencies toward physical activity in the relationship between behavioral apathy levels and the engagement in physical activity.

## 1. Introduction

Behavioral apathy is defined as a state of primary motivational impairment characterized by difficulty in elaborating the action plan required for behavior (Marin, 1990; Marin, 1991; Levy & Dubois, 2006). Apathy is observed in a wide range of conditions, including dementia (Leung et al., 2021), Parkinson’s disease (den Brok et al., 2015), and stroke (Zhang et al., 2023), and has been associated with frailty (Mega et al., 1996), functional decline (Ayers et al., 2017), poorer quality of life (Groeneweg-Koolhoven et al., 2014), higher mortality (Vilalta-Franch et al., 2013), and increased healthcare costs (Kruse et al., 2023). Additionally, higher levels of apathy have been associated with lower levels of physical activity (Farajzadeh et al., 2024). This association may further compromise the health of individuals with apathy, as insufficient physical activity is known to be associated with cognitive decline (Cheval et al., 2023), cardiovascular disease (Wahid et al., 2016), cancer (Moore et al., 2016), hypertension (Liu et al., 2017), diabetes (Cheval et al., 2021), obesity (Bleich et al., 2018), depression (Boisgontier et al., 2020), and functional dependence (van Allen et al., 2024a; van Allen et al., 2024b). Therefore, understanding the mechanisms underlying the association between behavioral apathy and physical inactivity may contribute to improving the design and specificity of rehabilitation interventions, ultimately improving the condition of individuals with apathy. However, these mechanisms remain largely overlooked and poorly understood. Considering their close relationship with both motivation and physical activity, intentions, explicit attitudes, and approach-avoidance tendencies toward physical activity may play a pivotal role in the relationship between apathy and physical activity engagement.

Intention is thought to capture the motivational factors that influence a behavior and reflects the effort that individuals are willing to invest for the behavior to occur (Ajzen, 1987). This motivational construct is considered the most proximal antecedent of physical activity behavior (Ajzen, 1987; Biddle et al., 2007). Further, the role of intentions in physical activity engagement is highlighted in a meta-analysis showing that less than 5% of individuals engage in physical activity without intending to be physically active (Feil et al., 2023). However, while intention is necessary to engage in physical activity, it is not sufficient to guarantee this engagement (Rhodes & de Bruijn, 2013). This insufficiency is illustrated by meta-analyses showing that almost half of the individuals who intend to be physically active are unable to fulfill this intention (Rhodes & de Bruijn, 2013; Feil et al., 2023).

In recent years, dual-process models have been developed to address the limitations of social cognitive models in explaining physical activity behavior (Rhodes et al., 2019). These new idiosyncratic models assert that physical activity behavior is governed not only by reflective processes (e.g., intentions, explicit attitudes), but also by automatic processes (e.g., automatic attitudes) (Conroy & Berry, 2017; Brand & Ekkekakis, 2018; Cheval & Boisgontier, 2021; Cheval & Boisgontier, 2024). Reflective processes are typically described as slow, deliberative, inefficient (relying on higher brain functions), and conscious (Strack & Deutsch, 2004), and are typically assessed with questionnaires. However, reflective processes do not always exhibit all these characteristics simultaneously, as certain reflective responses can be efficient or partially automatic (Melnikoff & Bargh, 2018). In contrast, automatic processes are faster and initiated unintentionally, rely on learned associations, do not require conscious awareness, and are most often assessed using reaction times to visual stimuli (e.g., images depicting physical activity vs. sedentary behavior).

Attitude is a psychological tendency to evaluate a stimulus with some degree of favor or unfavor (Eagly & Chaiken, 1993, p. 1). This tendency is considered an indirect antecedent of physical activity (Brand & Ekkekakis, 2018; Cheval & Boisgontier, 2021; Hagger & Chatzisarantis, 2014), influencing this behavior primarily through its effect on intention (Ajzen, 2011). Attitude can manifest as reflective (i.e., explicit attitudes) and automatic processes (i.e., automatic attitudes). Explicit attitudes are attitudes that people can report and for which activation can be consciously controlled (Rydell & McConnell, 2006). Automatic attitudes are introspectively unidentified traces of past experience that mediate favorable or unfavorable evaluation of a social object (Greenwald & Banaji, 1995). In other words, an automatic attitude is thought to result from the positive or negative value that our brain spontaneously assigns to some concept (e.g., person, place, or behavior), without that value being accurately accessible to cognition (Conroy & Berry, 2017). This implicit value of a stimulus results in an automatic positive or negative inclination toward this stimulus, which influences behavior. An illustration of this influence is approach-avoidance tendencies, an automatic preparation of the organism to execute a motor pattern toward or away from a behavior (Friese et al., 2011).

The aim of this study was to examine the relationships between apathy, the intention to be physically active, explicit attitudes toward physical activity, approach-avoidance tendencies toward physical activity and sedentary behaviors, and weekly physical activity engagement. According to a recent meta-analysis (Farajzadeh et al., 2024), we expected behavioral apathy to influence habitual levels of physical activity. Intention is essential for goal-directed behavior (Gollwitzer, 1997) and a key determinant of physical activity (Ajzen, 1987). Individuals with apathy often have difficulties initiating and maintaining goal-directed actions (Marin et al., 1991, Levy & Dubois, 2006). Therefore, we hypothesized that intention to engage in physical activity would mediate the relationship between apathy and habitual weekly physical activity engagement. The heightened sensitivity to effort-related cues in individuals with apathy leads them to perceive actions as more costly (Bonnelle et al., 2015; Bonnelle et al., 2016) and is likely to influence explicit attitudes toward physical activity as well as their antecedents, approach-avoidance tendencies. Therefore, our secondary hypothesis was that the relationship between apathy and intentions to be physically active would be mediated by explicit attitudes and approach-avoidance tendencies.

## 2. Methods

### 2.1. Population

Participants were recruited through social media, posters at the Faculty of Health Sciences, University of Ottawa, and emails to non-profit associations. Inclusion criteria were age 20–90 years and access to a personal computer, a laptop, or a tablet with internet. Informed consent was collected in accordance with the Declaration of Helsinki. The study was approved by University of Ottawa’s Research Ethics Boards (H-05-21-6791). All participants provided informed consent. Data were collected between July 2022 and December 2023. Participants were not compensated for their participation.

### 2.2. Power Analysis

An a priori power analysis was conducted in G*power (Faul et al., 2009) to estimate the minimum sample required for α = 0.05, power (1-β) = 90%, and a medium effect size f^2^ = 0.2 (Cohen, 1988). We used this heuristic value because, at the time the study was designed, no meta-analysis had assessed the effect size of apathy on physical activity or the effect of apathy on the intention to be physically active. The power analysis was based on an F test in a multiple linear regression (R^2^ increase) that included the two tested predictors (apathy and approach-avoidance tendencies) and 7 control variables and estimated that a minimum sample size of n = 67 was required. The aim of this approach was to ensure that we would have a 90% or greater chance of finding a statistically significant medium effect, if one exists, in each of the paths (i.e., each linear multiple regression model; Yzerbyt et al., 2018) of the mediation we described in our main hypothesis. However, because heuristics are a weak justification for determining an effect of interest, we aimed to collect as much data as possible within our time and resource constraints (Lakens, 2022). Of note, the meta-analysis by Farajzadeh et al. (2024), published after the data collection for the current study, provided two effect size estimates for the relationship between apathy and physical activity engagement: r = -0.13 (f^2^ = 0.017, based on r values in the literature) and r = -0.40 (f^2^ = 0.191, based on rho values in the literature). Using these effect sizes, analyses based on an F test in a multiple linear regression that included one tested predictor (apathy) estimated that a minimum sample size of n = 58 to 621 would have been required. Another approach would have been to use the powerMediation R package (Qiu, 2022). However, we decided against this approach because the lack of information available in the literature would have forced us to use heuristics to set the values of several arguments required for the power analysis.

### 2.3. Experimental Protocol

#### 2.3.1. Procedures

Participants performed approach-avoidance tasks online using Inquisit 6 software (Millisecond Software, 2015), and responded to questions related to apathy, habitual level of moderate-to-vigorous physical activity, age, sex (male, female), gender (man, woman, non-binary, transgender man, transgender woman, other), weight, height, depressive symptoms, and health condition. One attention check question was included in the questionnaires: “Please answer “5” to this question that allows us to verify that you actually read the questions.”

#### 2.3.2. Self-Reported Variables

##### Behavioral Apathy

Behavioral apathy was assessed using the action initiation subscale of the Lille Apathy Rating Scale (LARS) (Sockeel et al., 2006; Bonnelle et al., 2015). This subscale focuses on everyday productivity and initiative, and has been used as an index of behavioral apathy (Bonnelle et al., 2016). This subscale includes 11 questions with responses ranging from 1 (completely untrue) to 7 (completely true), resulting in a total score ranging from 11 to 77. We used the total score in our analyses, with higher scores indicating a higher level of behavioral apathy.

##### Habitual Level of Moderate-to-Vigorous Physical Activity

The habitual level of physical activity was derived from the short form of the International Physical Activity Questionnaire (IPAQ-SF), a self-administered questionnaire that identifies the frequency and duration of moderate and vigorous physical activity during the past 7 days to estimate habitual level of physical activity and sedentary behavior (Craig et al., 2003). The habitual level of moderate-to-vigorous physical activity (MVPA) in minutes per week was used in the analyses.

##### Explicit Attitudes

Explicit attitudes toward physical activity were calculated as the mean of two items based on two bipolar semantic differential adjectives on a 7-point scale [“unpleasant” (1) – “pleasant” (7); “unenjoyable” (1) – “enjoyable” (7)]. The statement begins with “For me, to participate in regular physical activity is …” (Hoyt et al., 2009; Farajzadeh et al., 2023). These two scores showed a very strong correlation (r = .915; p < 2.2×10^−16^), and their sum was used as an indicator of explicit attitudes in the statistical analyses.

##### Intention to Be Physically Active

The intention to be physically active was derived from the response to the question “How much do you agree with the following statement: Over the next 7 days, I intend to do at least 150 minutes of moderate-intensity physical activity; or at least 75 minutes of vigorous intensity physical activity; or an equivalent combination of moderate- and vigorous-intensity physical activity” on a 7-point scale ranging from “strongly disagree” (1) to “strongly agree” (7) (Rhodes & Rebar, 2017).

##### Depressive Symptoms

As it has been shown to be associated with apathy (Starkstein et al., 2001), depression needed to be controlled for in our analyses. Depressive symptoms were assessed using the Depression subscale of the Depression Anxiety Stress Scale (Lovibond & Lovibond, 1995). Participants were asked to read 14 statements and indicate how much the statement applied to them over the past week using a 4-point Likert scale ranging from 0 (“did not apply to me at all”) to 3 (“applied to me very much of the time”), resulting in a total score ranging from 0 to 48. We used the total score as a control variable in our analyses, with higher scores indicating greater depressive symptoms.

##### Chronic Conditions

Since it has been shown to be related with physical activity (Vancampfort et al., 2017), our analyses included the number of chronic conditions, which was derived from a question based on item PH006 of the Survey of Health, Ageing and Retirement in Europe (Börsch-Supan, 2022). “Has a doctor ever told you that you had any of the following conditions?”. The possible answers were “A stroke or cerebral vascular disease”, “High blood pressure or hypertension”, “High blood cholesterol”, “Diabetes or high blood sugar”, “arthritis, including osteoarthritis, or rheumatism”, “rheumatoid arthritis”, “chronic lung disease such as chronic bronchitis or emphysema”, “asthma”, “osteoporosis”, “Cancer or malignant tumour, including leukaemia or lymphoma, but excluding minor skin cancers”, “stomach or duodental ulcer, peptic ulcer”, “Parkinson disease”, “hip fracture or femoral fracture”, “Alzheimer’s disease, dementia, organic brain syndrome, senility or any other serious memory impairment”, “other affective or emotional disorders, including anxiety, nervous or psychiatric problems”, “chronic kidney disease”, “other conditions, not yet mentioned”, and “none”.

#### 2.3.3. Approach-Avoidance Tendencies

##### Approach-Avoidance Task

Approach-avoidance tendencies toward physical activity and sedentary stimuli were tested using an approach-avoidance task, which has shown good reliability (r = 0.83) (Farajzadeh et al., 2023) and good validity (Zenko & Ekkekakis, 2019). A video illustrating the task is available online (Boisgontier, 2024). Two experimental conditions and two neutral conditions were tested (Cheval et al., 2018; Farajzadeh et al., 2023). In the experimental conditions of this task, a trial starts with a fixation of a cross presented at the center of the screen for a random time ranging from 500 to 750 ms (Figure 1A). Then, an avatar appears either at the top or bottom third of the screen for one second, before a pictogram representing a physical activity behavior or a sedentary behavior appears in the center of the screen (Figure 1A). The participant sitting in front of the computer with one index finger positioned on the “U” and the other index finger on the “N” key is instructed that pressing the “U” key moves the avatar up and pressing the “N” key moves the avatar down. Accordingly, the movement of the avatar is always congruent with the pressed key: The top key (i.e., U) moves the avatar up, while the bottom key (i.e., N), moves the avatar down. Importantly, however, the approach or avoidance action depends on the initial position of the avatar at the beginning of the trial. If the avatar appears below the stimulus, the top key is associated with an approach movement, while the bottom key is associated with an avoidance movement. Conversely, if the avatar appears above the stimulus, the approach and avoidance movement are reversed – the top key is associated with an avoidance movement and the bottom key is associated with an approach movement.

**Figure 1.**
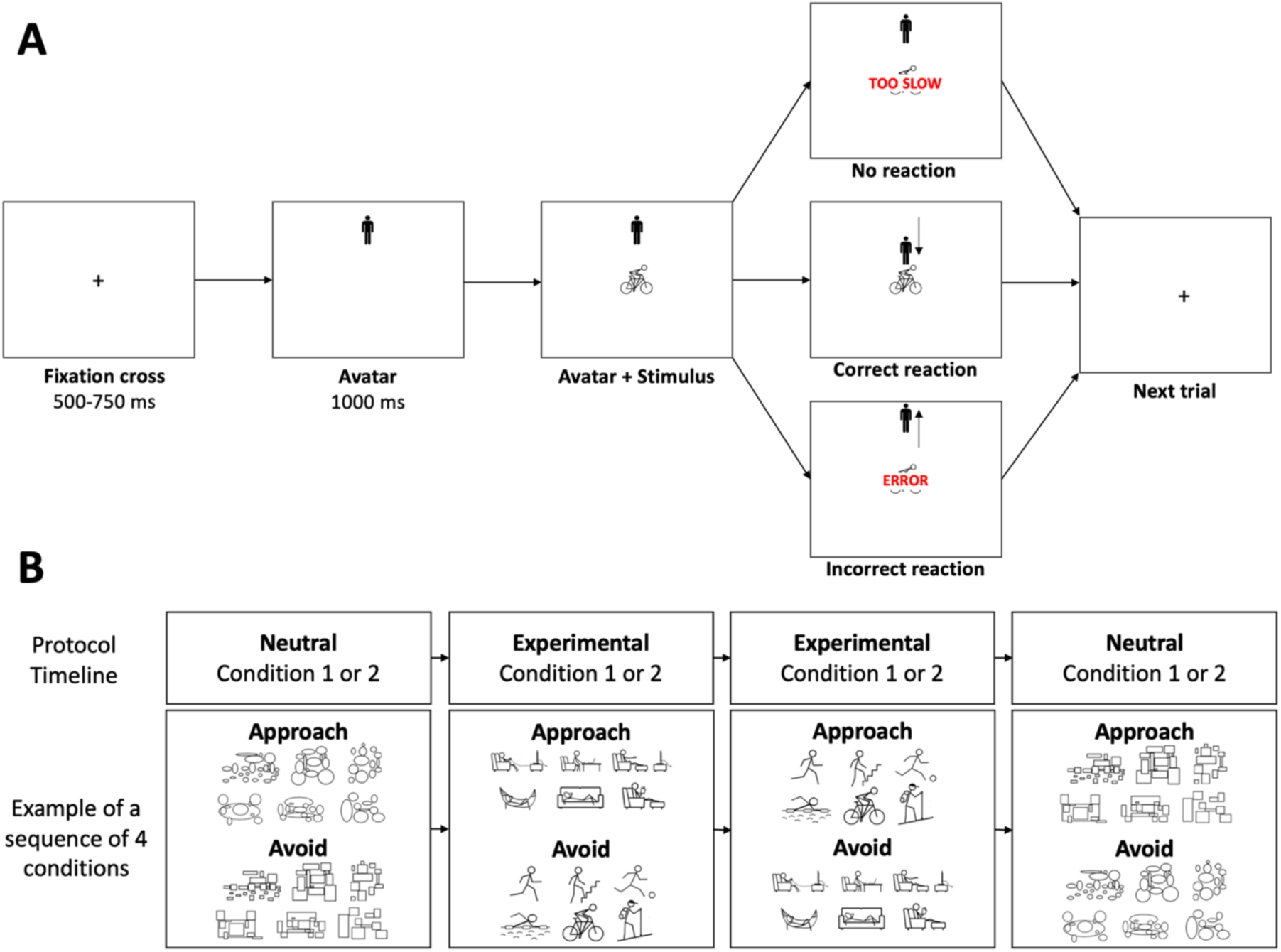
A. Illustration of a trial of the approach-avoidance task in the condition where the participant is instructed to approach physical activity stimuli (and avoid sedentary stimuli – not shown). B. Timeline and stimuli of the approach-avoidance task. In the experimental and neutral Condition 1, the participant is instructed to move the avatar toward (i.e., approach) a type of stimuli (i.e., physical activity or rectangles) and to move the avatar away from (i.e., avoid) stimuli depicting the other type of stimuli (i.e., sedentary behavior or ellipses, respectively). In Condition 2, the instruction is reversed: The participant is instructed to move away from physical activity (experimental condition) or rectangle stimuli (neutral condition) and move toward sedentary stimuli or ellipse stimuli.

##### Experimental Conditions

Two experimental conditions were tested (Figure 1B). In one experimental condition, the participant is instructed to quickly move the avatar toward (i.e., approach) pictograms depicting physical activity and to move the avatar away from (i.e., avoid) pictograms depicting sedentary behavior. In the other experimental condition, the participant does the opposite: move away from physical activity and move toward sedentary stimuli. The order of the experimental conditions was randomized across participants. In a previous study (Cheval et al., 2018), thirty-two participants were asked to rate the extent to which 24 stimuli expressed “movement and an active lifestyle” and “rest and sedentary lifestyle” (1 = not at all, 7 = a lot). For each stimulus, the “rest and sedentary lifestyle” score was subtracted from the “movement and active lifestyle” score. In the current study, the six stimuli with the largest positive and negative differences were chosen as the stimuli depicting physical activity and sedentary behaviors, respectively.

##### Neutral Conditions

In addition to the two experimental conditions, two neutral conditions were tested. These conditions were used to account for a potential generic approach-avoidance tendency that could vary across participants and ages (Farajzadeh et al., 2023). In these neutral conditions, the stimuli depicting physical activity and sedentary behaviors were replaced by stimuli made of rectangles or ellipses that matched the number and size of information in 3 physical activity stimuli (swimming, hiking, cycling) and 3 sedentary stimuli (couch, hammock, reading). Two conditions were tested. In one condition, participants are asked to quickly move the manikin toward stimuli with circles and away from stimuli with squares. In the other condition, the participant is given opposite instructions. The order of the neutral conditions was randomized.

One neutral condition was tested before the two experimental conditions, and the other neutral condition was tested after them. Each condition included 96 stimuli, 48 of each class (physical activity and sedentary stimuli in the experimental conditions; rectangles and ellipses in the neutral conditions), that were presented randomly. Familiarization with the task was performed during the first 15 trials of the study, which were removed from the analyses. Familiarization with the subsequent conditions was performed during the first 3 trials of each condition, which were removed from the analyses. The physical activity and sedentary stimuli were presented all together on the screen for seven seconds before each experimental condition. Between conditions, the participant could rest for as long as they wanted before pressing the space key to start the next condition. When the participant pressed the incorrect key (“U” when it should be “N” or “N” when it should be “U”), the message “error” appeared on the screen for 800 ms before the next trial. When the reaction time (i.e., the time between the appearance of the stimuli and the key press) was longer than eight seconds, the message “too slow” appeared on the screen for 800 ms before the next trial (Figure 1A).

##### Reaction Times

The tendency to approach or avoid a type of stimuli (i.e., physical activity, sedentary, or neutral stimuli) was derived from the time required to press the key in reaction to a type of stimulus (i.e., physical activity vs. sedentary vs. neutral). Corrected reaction times were computed by subtracting the mean reaction time to approach or avoid neutral stimuli from the reaction time on each trial to respectively approach or avoid stimuli depicting a type of behavior (physical activity or sedentary behavior). The bias toward a type of stimulus (physical activity or sedentary behavior) was computed by subtracting the mean corrected reaction time to approach this type of stimulus from the corrected reaction time to avoid it. Therefore, positive scores were indicative of an approach bias, i.e., a higher tendency to approach the stimulus. Incorrect responses, responses faster than 150 ms, and responses slower than 3,000 ms were excluded from the analyses to account for outliers and loss of attention (Farajzadeh et al., 2023). Specifically, because simple reaction times are typically longer than 200 ms (Fozard et al., 1994), and because we were assessing choice reaction times, we considered that reaction times faster than 150 ms could not be considered reactions to the stimuli of our task. In young adults, complex choice reaction times with up to 15 choices and different combinations of limb responses are typically faster than 1,000 ms (Boisgontier et al., 2014). However, in older adults, they can be longer than 3,000 ms (Boisgontier et al., 2016). Therefore, as with the lower threshold (150 ms), we chose a conservative upper threshold to avoid excluding a large proportion of valid reaction times in the older participants, as these observations can have a significant impact on the results (Ulrich & Miller, 1994; Miller, 2023). We did not include reaction times longer than 3000 ms because even in the complex task mentioned above (Boisgontier et al., 2016), the proportion of reaction times longer than 3,000 ms was less than 0.5% of the data. Therefore, we considered reaction times longer than 3,000 ms to be too long to be solely representative of a response to the stimuli of our task.

### 2.4. Statistical Analyses

To examine the mediation effect of intentions on the association between apathy and the habitual level of moderate-to-vigorous physical activity, we used the component approach (Yzerbyt et al., 2018). This component approach to the assessment of mediation was preferred over the index approach (Hayes, 2022), because the latter has shown a higher risk of false positives (Type I errors) (Yzerbyt et al., 2018). The component approach involves three linear multiple regressions models. Model 1 examines whether the independent variable affects the dependent variable. Model 2 examines the effect of the independent variable on the mediator. Model 3 examines both the independent variable and the mediator as simultaneous predictors of the dependent variable. Mediation was claimed if the “total effect” of the dependent variable in Model 1 is larger in absolute value than its “residual effect” in Model 3. The bias to approach physical activity stimuli, the bias to approach sedentary stimuli, depressive symptoms, age, sex, body mass index, and the number of chronic health conditions were included as control variable in all the models used to test the mediation. The first mediation analysis examined the intention to be active as a mediator of the effect of apathy on the habitual level of physical activity. The second mediation analysis examined explicit attitudes as a mediator of the effect of apathy on the intention to be active. The third mediation analysis examined approach-avoidance tendencies as a mediator of the effect of apathy on the intention to be active.

To further examine the effect of apathy on approach-avoidance attitudes related toward physical activity and sedentary stimuli, a linear and a logistic mixed-effect models (Lohse et al., 2023) were built and fit by maximum likelihood in the R software environment (R Core Team, 2023), using the lme4 (Bates et al., 2021) and lmerTest package (Kuznetsova et al., 2022), which approximates *p*-values using Satterthwaite’s degrees of freedom method. Continuous variables were standardized. For the linear mixed-effects model, restricted maximum likelihood (REML) was used as it provides less biased estimates of variance components than full maximum likelihood (Luke, 2017). Fixed effects included a three-way interaction effect of apathy (continuous), stimulus (physical activity vs. sedentary behavior), and action direction (approach vs. avoid) on corrected reaction time. The other fixed effects controlled for the effect of depressive symptoms, the habitual level of moderate-to-vigorous physical activity, age, sex, body mass index, the number of chronic conditions, and the device used to complete the study (computer vs. tablet). Our balanced design was fully crossed: Each participant was tested in the approach and avoid condition of four types of stimuli (physical activity, sedentary behavior, rectangles, ellipses), with each type including 6 pictograms. Therefore, we intended to include the random effect of participant, action direction, stimulus, and pictogram (Lohse et al., 2023). However, the model converged only when the random effects of stimulus was removed.

To ensure that the results obtained with the corrected reaction times cannot be explained by the speed-accuracy trade-off (Hick, 1952), we conducted a logistic mixed-effects model with the number of errors in the Approach-Avoidance Task as outcome. The structure of this model was similar to the linear mixed-effects models using reaction time as outcome. However, because the logistic mixed-effects models did not converge when the three-way interaction was included, we conducted a model with a two-way interaction between apathy and action direction on errors when reacting to physical activity stimuli and another model with the same interaction but in the conditions with sedentary stimuli. In addition, only the fixed effects of depressive symptoms, age, and explicit attitudes as well as the random effect of participant were included to allow for model convergence.

## 3. Results

### 3.1. Descriptive Results

#### 3.1.1. Participants

Three hundred and ninety two participants initiated the study. Some of them were included in a previous study from our group (Farajzadeh et al., 2022). Twenty three were excluded because they stopped the session before completing the study. Four participants were excluded because they answered the check question incorrectly. When participants reported height <50 cm or >250 cm or weight <30 kg or >250 kg), the data was removed and imputed using the sample mean. The final sample of 365 participants was 53.7 ± 17.9 years (mean ± standard deviation), with a mean score of behavioral apathy of 31.3 ± 9.3 out of a maximum of 77 (mean score per item of 2.9 ± 0.9 on a scale ranging from 1 to 7), a habitual level of moderate-to-vigorous physical activity of 398.3 ± 496.0 min per week, mean depressive symptoms of 0.7 ± 0.4, body mass index of 27.6 ± 12.1 kg.m^-2^, and 1.5 ± 1.5 chronic health conditions. Two males identified themselves as women. All the other male (n = 146) and female participants (n = 217) identified themselves as men and women, respectively.

**Table 1.** Descriptive statistics (n = 365, 148 females)

| **Variable** (minimum score-maximum score) | **Mean** | **SD** | **Range** |
| --- | --- | --- | --- |
| Age (years) | 53.7 | 17.9 | 20-89 |
| Body mass index (kg.m<sup>-2</sup>) | 27.6 | 12.1 | 15.1-48.7 |
| Moderate-to-vigorous physical activity (min.week<sup>-1</sup>) | 398.4 | 496.0 | 0-3080 |
| Explicit Attitude (2-14) | 11.0 | 3.2 | 2-14 |
| Intention (1-7) | 5.1 | 2.1 | 1-7 |
| Behavioral Apathy (11-77) | 31.3 | 9.3 | 11-60 |
| Depression (0-48) | 9.3 | 6.2 | 0-37 |
| Chronic Diseases (0-18) | 1.5 | 0 | 7 |
| Approach physical activity stimuli (ms) | 1094 | 547 | 163-2996 |
| Avoid physical activity stimuli (ms) | 1183 | 538 | 172-2998 |
| Approach sedentary stimuli(ms) | 1202 | 590 | 174-2996 |
| Avoid sedentary stimuli (ms) | 1153 | 535 | 192-2989 |
| Approach neutral stimuli (ms) | 1021 | 542 | 156-2999 |
| Avoid neutral stimuli (ms) | 1046 | 523 | 154-2997 |

#### 3.1.2. Cut-off score for behavioral apathy

In our study, 126 participants reported having no chronic condition. In this subsample, the mean score per item was 2.93 on a scale of 1 to 7, with a 95% confidence interval (95CI) ranging from 2.79 to 3.07. This result suggested that a mean score per item > 34.5% [(3.07 − 1) / (7 − 1)] of the scale range is indicative of behavioral apathy. Based on this cut-off score, participants with a score ≥ 34 would be classified as adults with apathy (i.e., 37.5% of the participants). The same approach applied to our full sample (n = 365) yielded similar results (95CI upper bound = 2.98; 33.1% of the scale range).

#### 3.1.3. Observations in the approach-avoidance task

A total of 71,378 trials were performed by the participants (Figure 2), of which 12,350 were excluded: 531 trials were aborted at eight seconds with the “too slow” message appearing on the screen, 768 were removed because the participants failed the attention check question, 3,965 reactions were longer than 3,000 ms and 119 were faster than 150 ms, and 6,967 were familiarization trials. Among the 59,028 trials included in the analyses, 55,050 were in the correct direction (14,523 for sedentary stimuli and 14,724 for physical activity stimuli, 25,803 for neutral stimuli). An error occurred in 3,978 trials. A total of 15,539 trials were reactions to sedentary stimuli and 15,662 to physical activity stimuli (including reactions in the correct and incorrect direction).”

**Figure 2.**
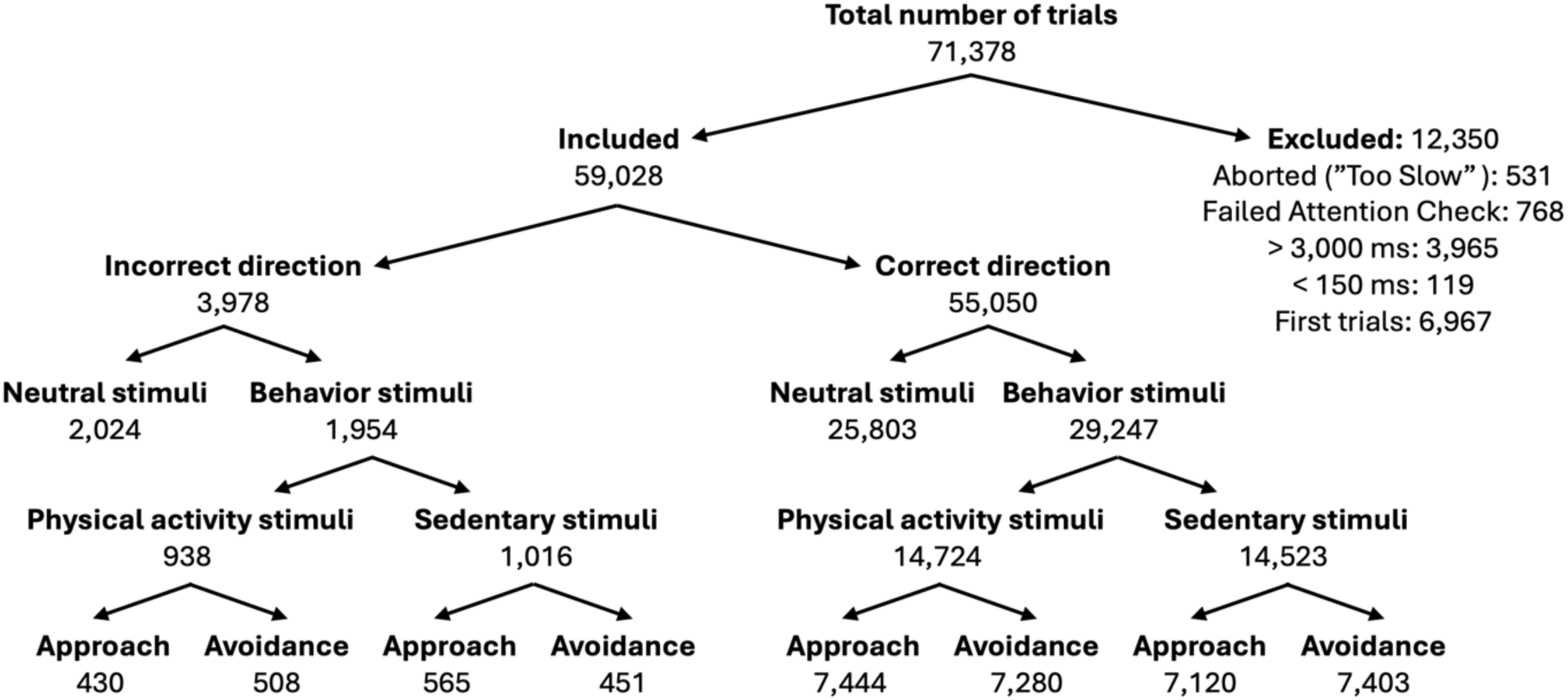
Flow diagram depicting the distribution of the trials

### 3.2. Statistical Results

#### 3.2.1. Intentions Mediate the Effect of Apathy on Physical Activity Engagement

Model 1 showed an association between apathy and the habitual level of physical activity (b = -67.6; 95CI = -119.1 to -16.2; p = .010). Model 2 showed an association between apathy and the intention to be physically active (b = -0.220; 95CI = -0.431 to -0.009; p = .041). Model 3 showed an association between intentions and physical activity (b = 147.3; 95CI = 97.4 to 197.3; p = 1.48 × 10^-8^). Model 3 also showed that the association between apathy and the level of physical activity was reduced when intentions were added as a predictor (b = -51.9; 95CI = -101.4 to -2.3; p = .040). From model 1 to 3, this association decreased by 23.3%. Taken together, these results demonstrate that the intention to be physically active partially mediates the relationship between apathy and the habitual level of physical activity (Figure 3).

**Figure 3.**
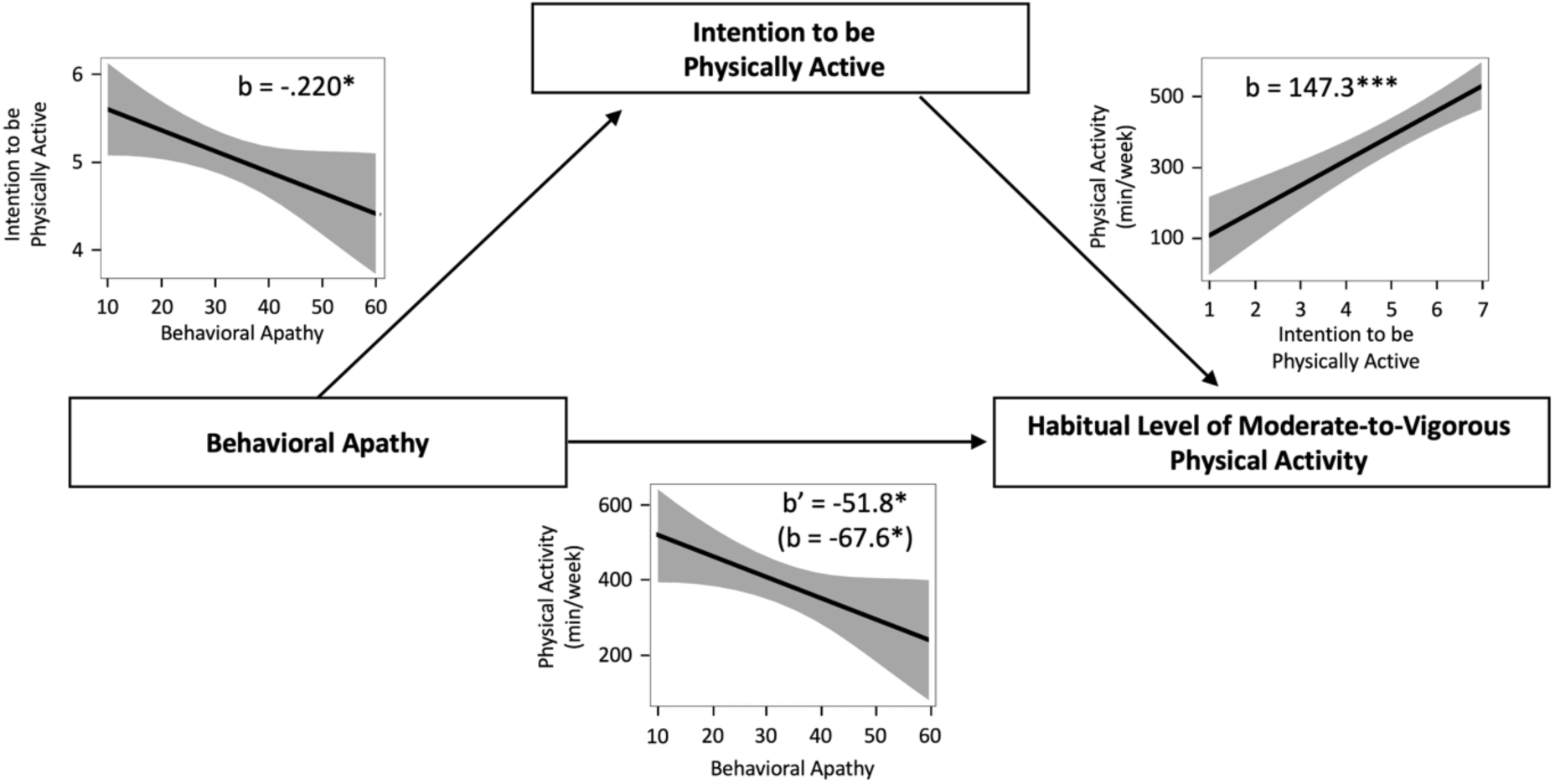
Mediation effect of intention to be physically active on the association between apathy and the habitual level of physical activity. Note: b’ is the estimate of the residual effect of apathy when intention is included in the model.

#### 3.2.2. Explicit attitudes Mediate the Effect of Apathy on Intentions

Model 1 showed an association between apathy and the intention to be physically active (b = -0.220; 95CI = -0.431 to -0.009; p = .041). Model 2 showed an association between apathy and explicit attitudes toward physical activity (b = -0.707; 95CI = -1.019 to -0.394; p = 1.15 × 10^-5^). Model 3 showed an association between explicit attitudes and the intention to physically active (b = 1.376; 95CI = 1.202 to 1.550; p < 2 × 10^-16^). Model 3 also showed that the association between apathy and intentions to be physically active was no longer significant when explicit attitudes were added as a predictor (b = 0.084; 95CI = -0.083 to 0.252; p = .324). Taken together, these results demontrate that explicit attitudes fully mediate the relationship between apathy and intentions to be physically active (Figure 4).

**Figure 4.**
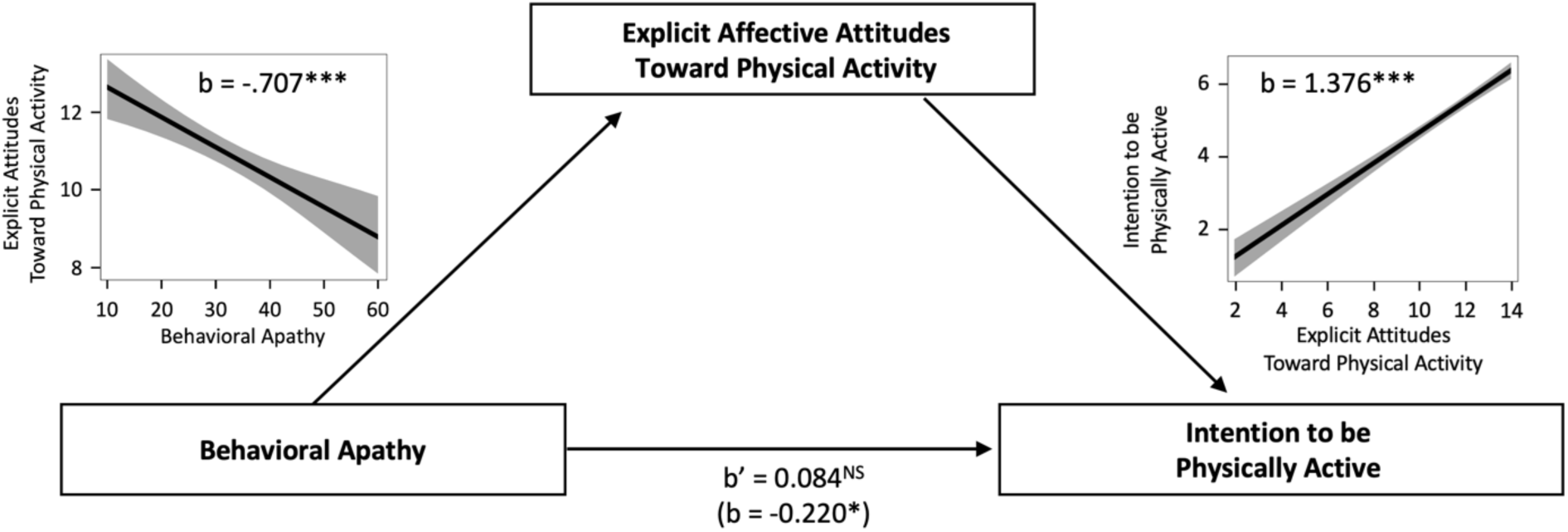
Mediation effect of explicit affective attitudes on the association between apathy and intentions to be physically active. Note: b’ is the estimate of the residual effect of apathy when explicit affective attitudes are included in the model. NS = non significant.

#### 3.2.3. Apathy and Approach-Avoidance Tendencies

Results of the linear mixed-effects model showed a significant three-way interaction between apathy (continuous), stimulus (physical activity vs. sedentary behavior), and action direction (approach vs. avoid) on corrected reaction time (b = 19.6; 95CI = 2.0 to 37.3; p = .029) (Figure 5). Simple effect analyses showed that lower apathy scores were associated with faster approach than avoidance of physical activity stimuli, but this difference was no longer significant when the apathy score was ≥ 55 out of a maximum mean score of 77. Additionaly, simple effect analyses showed that lower apathy scores were associated with faster avoidance than approach of sedentary stimuli, but this difference was no longer significant when the apathy scores were ≥ 70. In other words, lower apathy were associated with tendencies to approach physical activity stimuli and to avoid sedentary stimuli. Both these tendencies suggested a positive automatic evaluation of stimuli associated with physical activity. However, such tendencies decreased as apathy increased until they lost their statistical significance.

**Figure 5.**
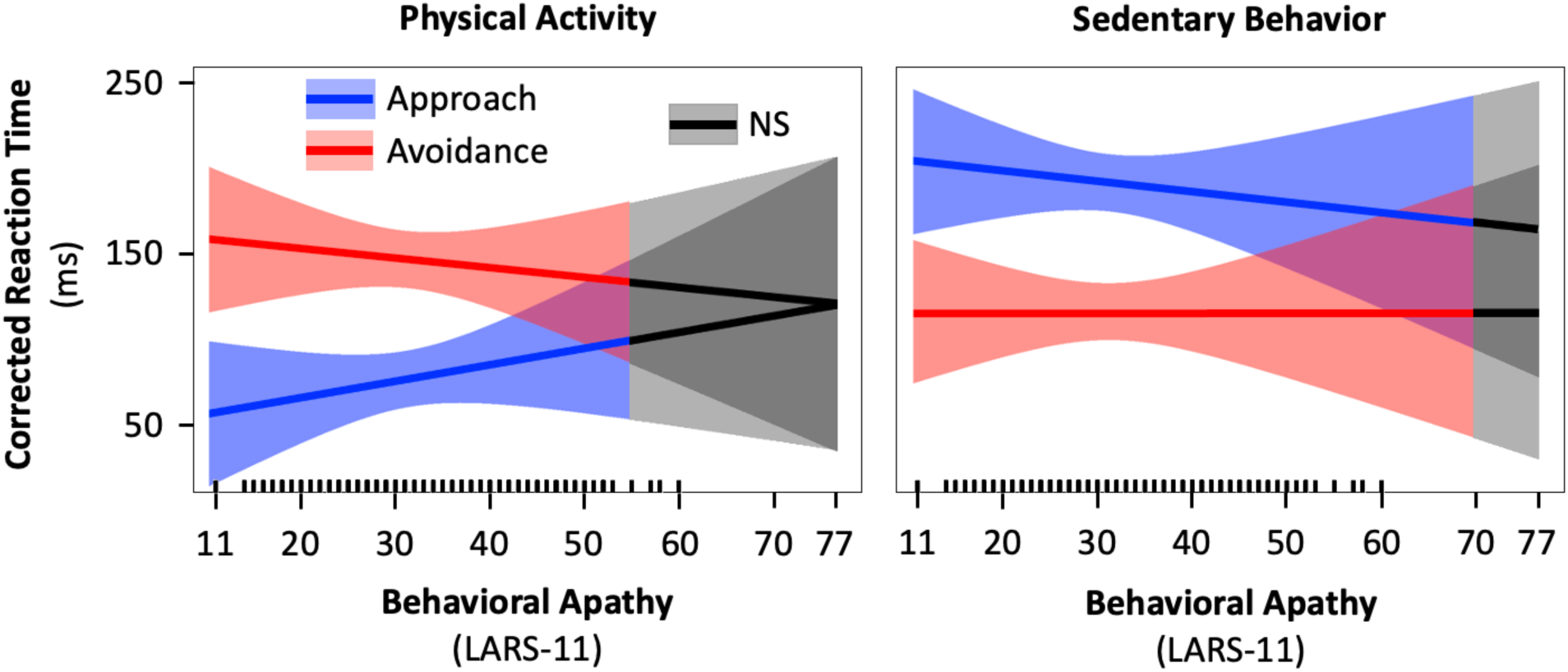
Results of the three-way interaction between the effect of apathy (continuous), stimulus (physical activity vs. sedentary behavior), and action direction (approach vs. avoid) on corrected reaction time. Note. LARS = Lille Apathy Rating Scale. NS = Non significant effect of action direction.

Results of the logistic mixed-effects models showed no evidence of an interaction between the effects of apathy and action direction on errors when reacting to physical activity (b = .135; 95CI = -.011 to .283; p = .07) or sedentary stimuli (b = .128; 95CI = -.025 to .283; p = .103). Additionally, results showed no evidence of an association between approach-avoidance tendencies and the habitual level of moderate-to-vigorous physical activity, ruling out the possibility for a mediation effect.

## 4. Discussion

### 4.1. Main findings

As hypothesized, weaker intention to be physically active mediated the association between higher behavioral apathy and lower habitual levels of moderate-to-vigorous physical activity. In addition, explicit attitudes mediated the relationship between behavioral apathy and intentions to be physically active. While higher apathy was associated with a stronger tendency to avoid physical activity stimuli and to approach sedentary stimuli, we found no evidence suggesting that this tendency mediated the relationship between apathy and intentions or habitual physical activity. Based on our data, a mean item score greater than 34.5% of the scale range (e.g., >3.07 on a 1–7 scale) is indicative of behavioral apathy.

### 4.2. Comparison with the Literature

#### 4.2.1. Behavioral Apathy Cut-Off Score

Our data in otherwise healthy older adults (n = 126) suggest that a mean score per item > 34.5% of the scale range is indicative of behavioral apathy. Based on this cut-off score, 37.5% of our participants (n = 365) would be classified as having behavioral apathy. Of note, this cut-off score was similar to the one based on our entire sample (33.1% of the scale range), which is consistent with the absence of statistical correlation between behavioral apathy and chronic conditions (r = -0.017, p = .742) and with the conceptualization of apathy as a syndrome rather than a symptom, since apathy levels were similar regardless of health status. In addition, our cut-off score is consistent with the mean item score of 29.7% of the scale range that was used as a cut-off score in another study of otherwise healthy young and middle-aged adults (n = 108, 94.5% aged 20 to 49 years), although they used a different scale, the Apathy Evaluation Scale ranging, which ranges from 1 to 4 (Kant et al., 1998). However, other results in otherwise healthy young adults (n = 139) showed a mean apathy score per item of 1.46 on a scale of 1 to 5, with an upper bound 95CI of 1.58 (Bonnelle et al., 2016). Accordingly, they considered a mean score per item of 14.5% of the scale range to reflect behavioral apathy. Based on this cut-off score, 88.5% of our sample would have been classified as adults with apathy. We initially hypothesized that this higher threshold for apathy in our study compared to the study by Bonnelle et al. (2016) might be due to the broader age distribution (mean age: 27.4 ± 7.8 vs. 53.7 ± 17.9 years) since a recent meta-analysis showed stronger correlation between apathy and physical activity in older adults compared to younger adults (Farajzadeh et al., 2024). However, our results showed no evidence of a positive correlation between behavioral apathy and age in the healthy subset (r = -0.173, p = .053).

#### 4.2.2. Behavioral Apathy and Physical Activity

The association between apathy and physical activity observed in the current study aligns with results of a recent meta-analysis, which reported a negative correlation based on 22 studies (n = 12,541) (Farajzadeh et al., 2024).

#### 4.2.3. Mediating Role of Intentions

For the first time, we show an association between behavioral apathy and intentions to be physically active. This finding suggests that behavioral apathy affects the intention stage of goal-directed behaviors (Levy & Dubois, 2006), such as physical activity. In addition, this finding is conceptually coherent because behavioral apathy, a motivational impairment (Marin, 1990; Marin, 1991; Levy & Dubois, 2006), is likely to reduce the amount of effort individuals are willing to invest for the physical activity behavior to occur. This reduction could be explained by an increased sensitivity to effort (Bonnelle et al., 2016) and/or a reduced sensitivity to rewards associated with physical activity (Pessiglione et al., 2018), both of which could be linked to changes in brain circuitry, even at subclinical levels of apathy (Derosiere et al., 2024).

Our results also show an association between the intention to be physically active and the engagement in physical activity behavior, which is in line with previous theoretical and experimental literature (Ajzen, 1987; Biddle et al., 2007; Maltagliati et al., 2024a). Recent experimental results based on the theory of effort minimization in physical activity (Cheval & Boisgontier, 2021; Cheval & Boisgontier, 2024) suggest that this association between intentions and physical activity behavior is influenced by individual differences in the general tendencies to approach and avoid physical effort (Cheval et al., 2024; Maltagliati et al., 2024a). Specifically, the positive association between intentions to be physically active and physical activity engagement was stronger when self-reported tendencies to approach physical effort were higher, and weaker when tendencies to avoid physical effort were higher. These findings encourage future studies to examine whether approach-avoidance tendencies toward physical effort influence the relationship between behavioral apathy and physical activity engagement.

The observed mediating effect of intention suggests that interventions aimed at increasing physical activity levels in patients with apathy should first focus on enhancing their intention to be physically active. While such intention does not guarantee engagement in physical activity, it is an essential prerequisite for this engagement (Rhodes & de Bruijn, 2013; Feil et al., 2023).

#### 4.2.4. Mediating Role of Explicit Attitudes

For the first time, we show an association between behavioral apathy levels and explicit affective attitudes toward physical activity, which mediate the relationship between apathy and intentions to be physically active. These attitudes are based on the aggregation of the likelihood of anticipated affective responses (Williams & Evans, 2014; Stevens et al., 2020). Therefore, to strengthen positive explicit attitudes in individuals with apathy, ensuring that exercise therapy is consistently associated with positive experiences may be important. This suggestion is consistent with existing literature highlighting the role of pleasure and displeasure in physical activity engagement (Ekkekakis, 2017; Maltagliati et al., 2024b). Ultimately, this effect of explicit attitudes on intentions may influence behavior as within-subject increases in positive affect have been shown to be associated with physical activity levels at 3-month follow-up (Kwan & Bryan, 2010).

#### 4.2.5. Physical Activity and Approach-Avoidance Tendencies

Literature has consistently reported a tendency to approach physical activity stimuli and to avoid sedentary stimuli at all ages (Farajzadeh et al., 2023). In the current study, we showed that these tendencies are influenced by apathy. Participants with stronger behavioral apathy showed weaker tendencies to approach physical activity and avoid sedentary behaviors. Approach-avoidance tendencies reflect motivational processes that direct organisms toward beneficial stimuli (approach) and away from detrimental stimuli (avoidance), driving adaptive behavior (Elliot et al., 2013). These tendencies are driven by internal states and action plans that translate perceived urges into behavior (Sander et al., 2018). Specifically, factors such as the perceived reward and the amount of effort involved in a behavior can influence whether a person is motivated to approach or avoid this behavior. Apathy, characterized by reduced motivation and difficulty generating action plans for goal-directed behavior, results from a desensitization to reward (Martínez-Horta et al., 2014; Muhammed et al., 2016) or an increased sensitivity to effort (Bonnelle, 2016; Le Heron, 2018; Le Heron, 2019). Therefore, individuals with apathy may perceive physical activity as more effortful and less rewarding than individuals without apathy. Thus, the threshold of the reward-to-effort ratio that triggers behavior may be harder to reach, reducing the likelihood of approaching physical activity. This example illustrates how apathy can disrupt the approach-avoidance dynamic and dampen the internal drive toward physical activity behavior.

### 4.3. Limitations and Strengths

The present study has potential limitations. First, the online nature of the study made it impossible to limit the influence of potential distractions in the participant’s environment and to control whether participants were using their two index fingers to perform the task as instructed and whether they were sitting or standing, which could have influenced the results (Cheval et al., 2018; Maltagliati et al., 2024c). Second, the participants were recruited in Canada. It is, thus, unclear whether conclusions could generalize to populations from non-Western countries or less active populations of older adults. Third, the habitual level of physical activity was assessed using a self-reported questionnaire, which may not accurately reflect the objective level of physical activity. Assessing physical activity and sedentary behaviors using device-based measures would have provided more reliable estimates. However, these limitations are offset by several strengths, including a large sample size, a measure that accounts for a potential generic approach-avoidance bias that could have confounded the results (Farajzadeh et al., 2023), and the use of statistics that limit information loss (i.e., mixed effects models) (Boisgontier & Cheval, 2016; Lohse et al., 2023).

## 5. Conclusion

This study supports previous work suggesting that apathy is a syndrome rather than a symptom (Marin et al., 1991), affecting not only individuals with a medical condition but also otherwise healthy adults across all ages. In addition, the results highlight the key role of intentions and explicit affective attitudes in mediating the relationship between behavioral apathy and habitual physical activity. These insights suggest that interventions aimed at improving or maintaining physical activity levels should assess apathy regardless of a patient’s or client’s health status. For individuals identified as having apathy, incorporating motivational constructs—such as the intention to be physically active and positive affective attitudes toward physical activity—into the intervention design should be considered.

## Data Availability

All data produced and code are available online at https://doi.org/10.5281/zenodo.12750404

## 6. Declarations

### Data and Code Availability

According to good research practices (Boisgontier, 2022), the dataset and the R script are available in Zenodo (Boisgontier, 2024).

### Authorship Contribution Statement

Based on the Contributor Roles Taxonomy (CRediT) (Allen et al., 2019), individual author contributions to this work are as follows: Ata Farajzadeh: Writing – Review and Editing. François Jabouille: Writing – Review and Editing. Nickolas Benoit: Investigation. Olivier Bezeau: Investigation. Tristan Bourgie: Investigation. Benjamin Gerro: Investigation. Jacob Ouimet: Investigation. Matthieu P. Boisgontier: Conceptualization (Lead); Methodology; Formal Analysis; Data Curation; Visualization; Writing – Original Draft; Writing – Review and Editing; Supervision (AF, FJ, NB, OB, TB, BG, JO); Project Administration; Funding Acquisition. All authors have read and agreed to the published version of the manuscript.

